# Assessing required SARS-CoV-2 blanket testing rates for possible control of the outbreak in the epicentre Lusaka province of Zambia with consideration for asymptomatic individuals: a simple mathematical modelling study

**DOI:** 10.1101/2020.07.12.20152124

**Authors:** Joseph Sichone, Musalula Sinkala, Mervis Kikonko, Sody M. Munsaka, Martin Simuunza

## Abstract

**Introduction:** The novel Coronavirus disease (COVID-19), caused by the severe acute respiratory syndrome coronavirus - 2 (SARS-CoV-2), in Africa is characterised by a more substantial proportion of asymptomatic (or mildly symptomatic) individuals thought to be playing a role in the spread of the infection. The exact proportion and degree of infectiousness of asymptomatic individuals remains unclear. Studies however indicate that their management is crucial for control of SARS-CoV-2 transmission.

**Methodology:** We developed a simplified deterministic susceptible-exposed-infectious-removed (SEIR) mathematical model to assess the effect of active isolation of SARS-CoV-2 infected but asymptomatic individuals through blanket testing for control of the outbreak in Lusaka Province of Zambia. Here we modelled two scenarios; (1) assuming asymptomatic individuals comprised 70% of all COVID-19 cases and (2) asymptomatic individuals comprised only 50% of the cases. For contrast, the model was assessed first under the assumption that asymptomatic individuals are equally as infectious as symptomatic individuals and then secondly, and more likely, assuming asymptomatic individuals are only half as infectious as symptomatic individuals.

**Results:** For the model assuming 70% asymptomatic cases, a minimum sustained blanket testing rate of ≥ 7911 tests/100000 population was sufficient to control the outbreak if asymptomatic individuals are only half as infectious while if equal infectiousness was assumed then a testing rate of ≥ 10028 tests/ 100000 population would be required. For 50% asymptomatic, minimum blanket testing rates of ≥ 4540 tests/ 100000 population was sufficient to control the outbreak at both assumed levels of infectiousness for asymptomatic individuals relative to symptomatic individuals.

**Discussion and conclusion:** Our model predicts that the current testing rates of ≈ 150/100,000 population are inadequate to control transmission of SARS-Cov-2 in Lusaka. Active isolation of COVID-19 cases including asymptomatic individuals through blanket testing can be used as a possible measure for control of the SARS-Cov-2 transmission in Lusaka, Zambia.

## Introduction

Since the first reported case on 31 December 2019 in China, the current pandemic of the novel Coronavirus disease (COVID-19), caused by severe acute respiratory syndrome coronavirus - 2 (SARS-CoV-2), has killed over 327,700 and infected over 4.9 million people globally by 21^st^ May, 2020 [52]. The disease is characterised by a more substantial proportion of asymptomatic (or mildly symptomatic) individuals thought to be playing an “in-dismissible” role in the spread of the infection [2,3,4,24,25,33,64]. The degree of infectiousness of the asymptomatic individuals remains unclear although recent data from the World Health Organisation (WHO) and others suggest that relative to individuals with symptomatic infections, those with asymptomatic infection are considerably less infectious. [57,61,62]. Nonetheless, various studies, including modelling studies, have suggested that asymptomatic individuals comprise about 50 - 80% of all COVID-19 cases and may be responsible for as much as 40 – 73% of new infections [3,4,53,55,56]. Therefore, their management is critical for control of the disease [3,4,33]. To give a striking example, identification and isolation of asymptomatic people through blanket testing helped eliminate the virus in a completely isolated village of about 3000 people in northern Italy which saw the number of people with COVID-19 symptoms fall by over 90% within ten days [3]. The current study applied a simple mathematical modelling approach to explore the effect of increased blanket testing rates as a possible measure to capture and isolate asymptomatic individuals and control the COVID-19 outbreak in Lusaka Province of Zambia which is the epicenter of the outbreak in Zambia since the first recorded case in the country on 18^th^ March 2020. [44]. Recent studies have modelled the spread and expected burden of the COVID-19 outbreak in Africa and Zambia and explored the effects of various control measures such as applying different levels of physical distancing and shielding in the population [42,43,51,72]. Although this provides vital information to guide policy for Zambia, some interventions may not be easy to monitor in practice. Additionally, while such interventions have already been instituted, cases continue to rise in Zambia and other African countries [20,43,44,50]. Assessment of more COVID-19 control options through mathematical modelling based on the known epidemiology of the disease would therefore serve to supplement current information on the possible management of the outbreak in Zambia.

## Materials and Methods

### Study Area

Lusaka Province is the smallest and highly urbanised province in Zambia (83.5% urbanisation) with seven districts over an area of about 23,490 km^2^ [45]. It is one of the most densely populated provinces in the country [46,47]. It has a total population of about 3,308,438 - Density: 140.8/km^2^ (2019) [45]. The provincial capital, which is also the capital of Zambia, is the highly-populated Lusaka district with the latest population estimated at 2,627,700 – Density: 6,288/km^2^ (2019) [45]. Lusaka is a busy corporate and commercial hub of Zambia and an outlet to the rest of the world with the busy Kenneth Kaunda International Airport. It is therefore no surprise that the first recorded COVID-19 cases in Zambia occurred in Lusaka district as imported cases [44]. The province also shares borders with neighboring Zimbabwe and Mozambique [47].

### Lusaka Province COVID-19 Outbreak

On 18^th^ March 2020, Zambia recorded the first confirmed cases of COVID-19 in Lusaka from two residents who had previously travelled to France [44]. By 21^st^ May 2020, Lusaka had recorded about 291 confirmed cases and 6 deaths [44]. From the onset of the outbreak in Lusaka, the Zambian Ministry of Health and other stakeholders had implemented required preparedness and disease control measures including mandatory physical distancing, surveillance and case notification, heightened sanitation and handwashing in public places, closure of some public institutions as well as general sensitisation [44]. The main laboratory testing approach employed has been targeted testing based on prescribed case definitions and contact tracing to optimise positivity rates (Zambia COVID-19 situation reports No. 1-14) [44]. A few mass testing campaigns have also been conducted in some areas of Lusaka province as by 21^st^ May 2020 e.g. Rhodes park (about 1,190 persons tested), Chirundu, (about 1,000 persons tested), and Kafue (undetermined) (Zambia COVID-19 situation reports No. 29,44-46,62-64) [44].

### The Model

The spread of SARS-Cov-2 in Lusaka province was modelled through a simplified deterministic susceptible-exposed-infectious-removed (SEIR) compartmental mathematical model as shown in Fig. 1.

**Figure 1.**
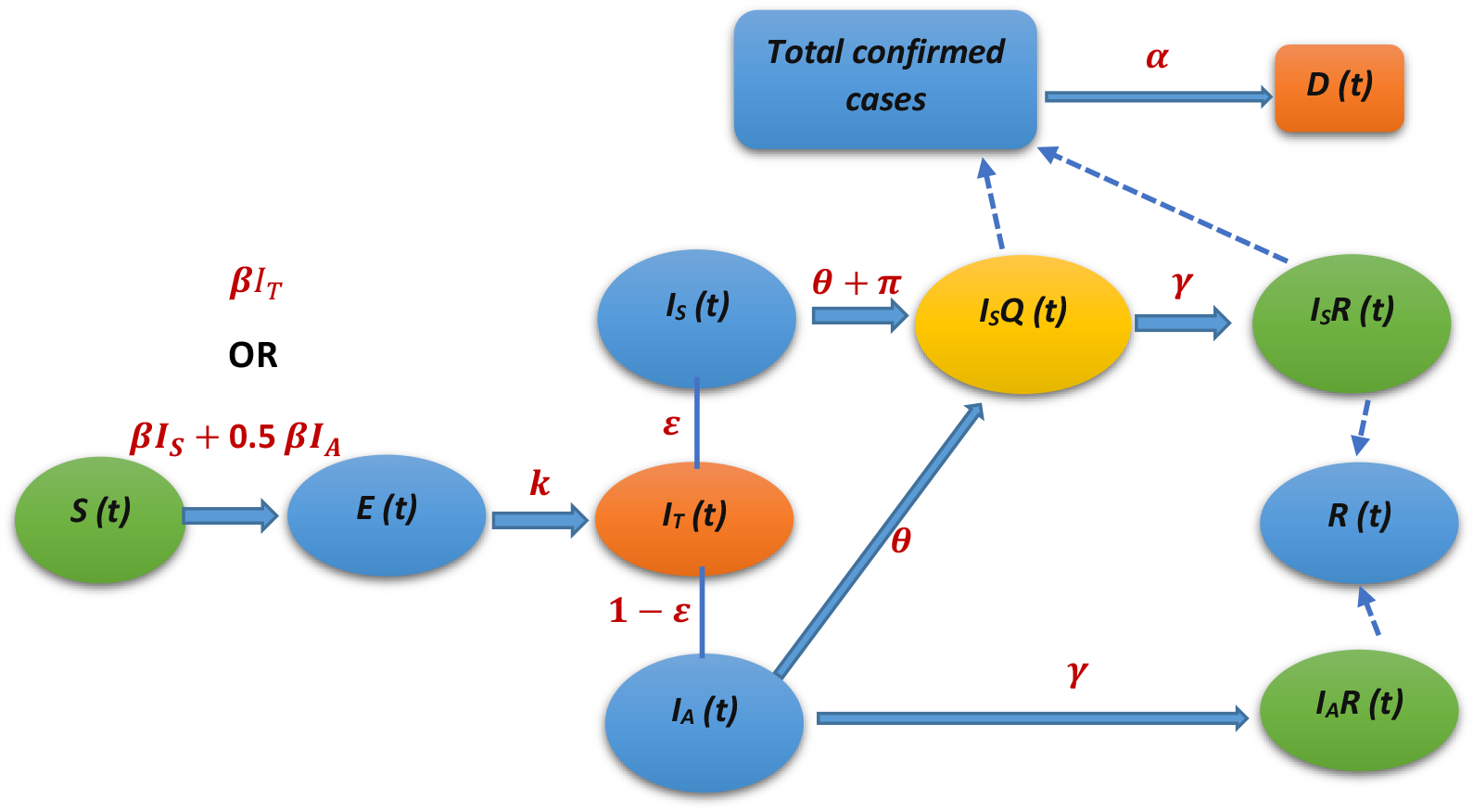
The SEIR model for spread of COVID-19 in Lusaka province. The solid arrows represent flow between compartments and dotted arrows represent additive contribution relationship to a compartment. Infectious individuals are split into symptomatic and asymptomatic individuals.

The model both directly and indirectly incorporated the current mitigation measures in place to attempt to predict the trajectory of the outbreak in Lusaka accurately. We started by denoting the infection states as total number of susceptible *S(t)*, exposed *E(t)*, infectious *I(t)*, removed *R(t)* and dead persons *D(t)* at any given time *(t)* in the population of size *N*. For our analysis, the total population size was assumed to be constant and demographics of natural birth and deaths rates were considered negligible [17,70]. Table 1 shows the average values of parameters used and equations in set 1 and set 2 give the systems of ordinary differential equations (ODEs) describing the flow of individuals in the model. The ODEs in set-1 describe spread of infection under the assumption of equal infectiousness for both asymptomatic and symptomatic individuals while the ODEs in set-2 describe the spread of infection assuming asymptomatic individuals are only half as infectious as symptomatic individuals. For the ODEs in set-1, we assume that susceptible individuals are infected at the rate 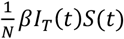 where *I*_*T*_(*t*) is the total number of active infectious individuals in the population both symptomatic *I*_*S*_(*t*) and asymptomatic *I*_*A*_(*t*). On the other hand, for the ODEs in set-2, the susceptible individuals are infected at the rate 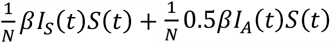 with the probability of infection from asymptomatic individuals only half that of symptomatic individuals. In both equations, after infection occurs the susceptible individuals are exposed *E*(*t*) and enter an incubation period of about 4.8 days before they become infectious *I*(*t*). Once they have become infectious, they may belong to either one of two classes of infectious individuals; symptomatic infectious (*I*_*S*_), or asymptomatic infectious (*I*_*A*_) determined by the fraction for symptomatic persons ɛ. Based on current interventions in Lusaka, the symptomatic infectious individuals are effectively managed in that once a person becomes symptomatic in the community, they are identified through targeted testing under the current testing criteria and quarantined. However, in our model we assumed an average delay of about 2.5 days in the identification of these symptomatic individuals because sometimes people delay in seeking medical attention when symptoms are unclear, may be in self-denial, plus have an extra 24hrs turnaround time delay in laboratory results [44]. The symptomatic infectious individuals *I*_*S*_*Q*(*t*) are therefore quarantined at a rate *π* (1/diagnosis delay). It is taken that during this delay period these symptomatic individuals can infect others but are no longer able to do so once isolated in quarantine. The quarantined infectious individuals recover at rate *γ* and are removed as *I*_*S*_*R*(*t*). On the other hand, in our model the asymptomatic (or mildly symptomatic) infectious individuals (*I*_*A*_) are taken to be generally unnoticed in the community but also recover at a rate *γ* in which time they can infect susceptible individuals before they become removed as *I*_*A*_*R*(*t*) and no longer infectious. Note that some asymptomatic individuals develop symptoms much later in their infection but this does not substantially affect our model because at that time they would still be removed (*I*_*A*_*R*) if they become diagnosed and quarantined through targeted testing. In this study, we assumed that the current mostly targeted testing for COVID-19 in Lusaka province is restrictive and probably missing some asymptomatic individuals [53]. Therefore, a parameter (*θ*) was introduced in the model which describes blanket testing applied as tests per 100000 populations used to identify and isolate all infectious individuals in the community (symptomatic and asymptomatic) through sustained random mass testing. The total removed individuals for the model *R*(*t*) are given as *I*_*S*_*R*(*t*) + *I*_*A*_*R*(*t*) while the total confirmed cases are given as *I*_*S*_*Q*(*t*) + *I*_*S*_*R*(*t*) of which a fraction *α* (Case fatality rate -CFR) are recorded dead *D*(*t*).

**Table 1.** Parameters used for the SEIR model for spread of COVID-19 in Lusaka Province

| Parameter | Definition | Value | Comment | Reference |
| --- | --- | --- | --- | --- |
| $N$ | Total population | 3300000 | Lusaka | [45] |
| $R_0$ | Basic reproduction number | - | Data fitted | [1,2,12,16,17,21,27,28,32,33,37] |
| $\beta$ | The average rate of effective contacts per unit time between susceptible people and infectious people | $\gamma R_0$ | Data fitted | - |
| $k$ | The average rate to infectiousness | 1/4.8 | (1/incubation period) | [1,2,5,6,8,16,17,21,27,28,33,35,39,41] |
| $\gamma$ | Recovery rate | 1/7.5 | (1/infectious period) | [1,2,5,8,14,16-19,21,24,26,27,33,35,39] |
| $\varepsilon$ | Fraction symptomatic infectious people | 30% - 50% | - | [3,4,24,33] |
| $\pi$ | Quarantine rate for symptomatic | 1/2.5 | (1/ diagnosis delay). Assumed. | [6] |
| $\theta$ | Community blanket testing rate | 150/100000 – 10733/100000 | (tests per 100000 population) | - |
| $\alpha$ | Apparent death rate (CFR) | 0.0206186 | Data fitted | - |

### Model optimisation and simulation

Model optimisation and simulation was done using Vensim PLE systems dynamics modelling software for Windows (version-7) [48]. This was done for two scenarios of 70% and a modest 50% assumed proportion of asymptomatic infectious individuals in the population. Data from the first three months of the outbreak in Lusaka as given in the Zambia COVID-19 situation reports No. 1-64 [44] was used to configure the model and optimise parameters. However, due to presence of imported cases in the early days and the considerable variations in recorded cases between some days (probably influenced by variations in availability of testing kits), only data from 10^th^ April 2020 to 16^th^ May 2020 was used. This is because this period had more consistent data and by then community infections had been established [44]. The model initial conditions were estimated from the available data as follows: *S (0)* = *N*-92, *E (0)* = 24, *I*_*S*_ *(0)* = 9, *I*_*S*_Q *(0)* = 38, *I*_*S*_*R (0)* = 0, *I*_*A*_ *(0)* = 21, *I*_*A*_*R (0)* = 0, *R (0)* = 0, and *D (0)* = 0. With other parameter values fixed, the model was calibrated to the cumulative number of confirmed cases over time. This was done by adjusting values of R_0_ until the best model fit was achieved (since R_0_ was expectedly affected by the current mitigation measures). R_0_ estimates the average number of secondary infections arising from a single infectious individual in a naive population. Model fit was statistically evaluated using Spearman correlation and chi-square goodness of fit test at a significance level of 0.05. For the calibration, *θ* was approximated at 150 tests/100000 populations – reflective of the mass testing rates achieved in this period [44]. After calibration, the model simulation was extended to 631 days (10^th^ April 2020 – 31^st^ December 2021) to predict the spread of the outbreak in Lusaka under the current transmission rate. To assess which value of *θ* would sufficiently flatten the curves of both the total number of active infectious individuals and the cumulative number of confirmed cases over time (as a key indicator of control of the outbreak), several iterations of this simulation were then performed using increasingly higher values for *θ*.

### Ethical Considerations

No ethical issues were encountered as no human or animal subjects were used in this study and cases were anonymous.

## Results

The model had significant fit to outbreak data under all the assessed conditions and therefore could be used for the general purpose of analysing the outbreak under all these general scenarios. Figure 2 shows results of model optimisation and fit to outbreak data for both the 70% and 50% asymptomatic scenarios assuming equal infectiousness for asymptomatic and symptomatic individuals (denoted as A1 and A2). Figure 3 shows the model fit results when asymptomatic individuals are only half as infectious as symptomatic individuals (B1 and B2). All R_0_ values were within the range estimated for COVID-19 [1,2,12,16,17,21,27,28,32,33,37].

**Figure 2.**
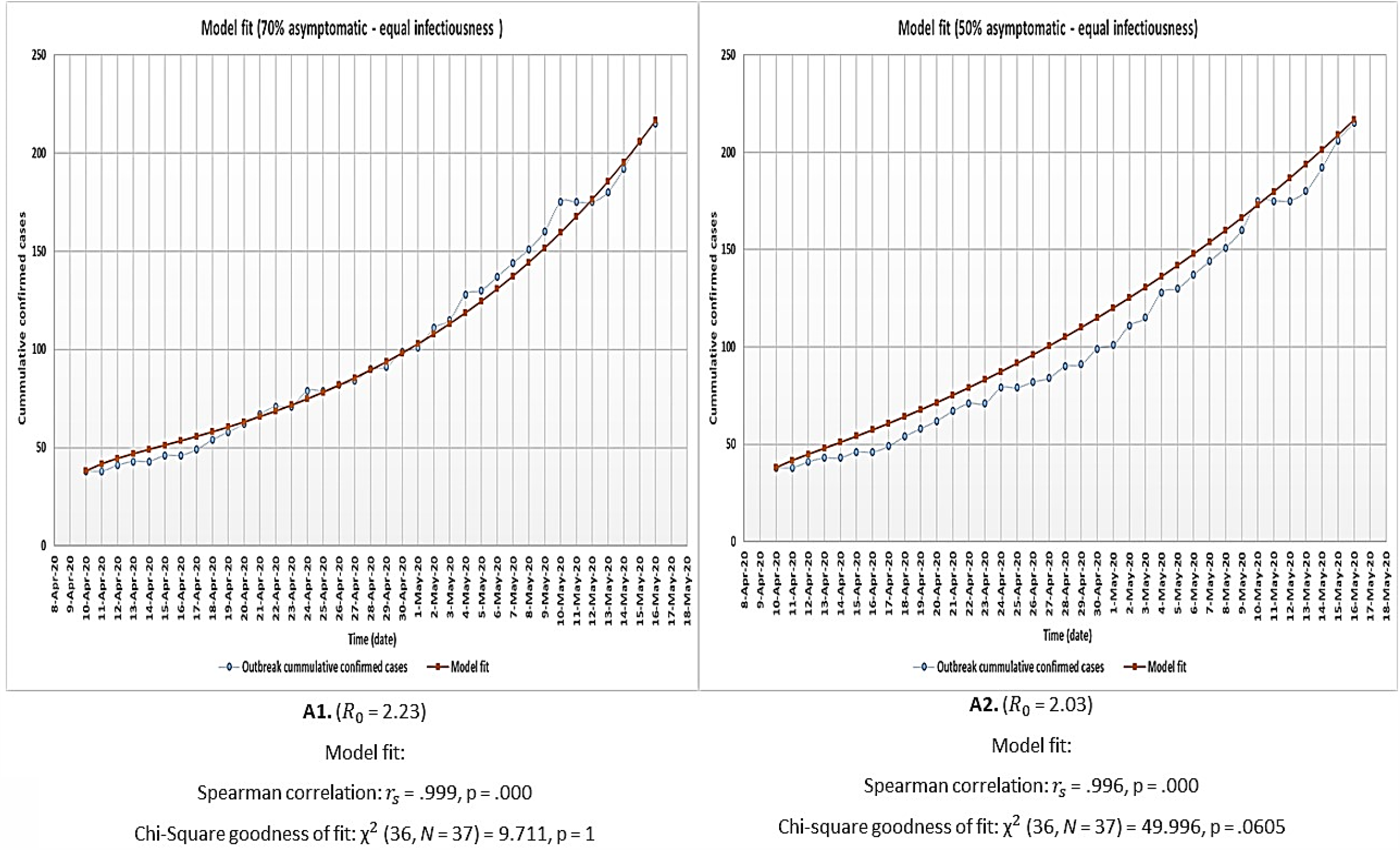
Model optimisation and fit to outbreak data for both the 70% (A1) and 50% (A2) proportion asymptomatic individual’s scenarios under the assumption of equal infectiousness for asymptomatic and symptomatic individuals.

**Figure 3.**
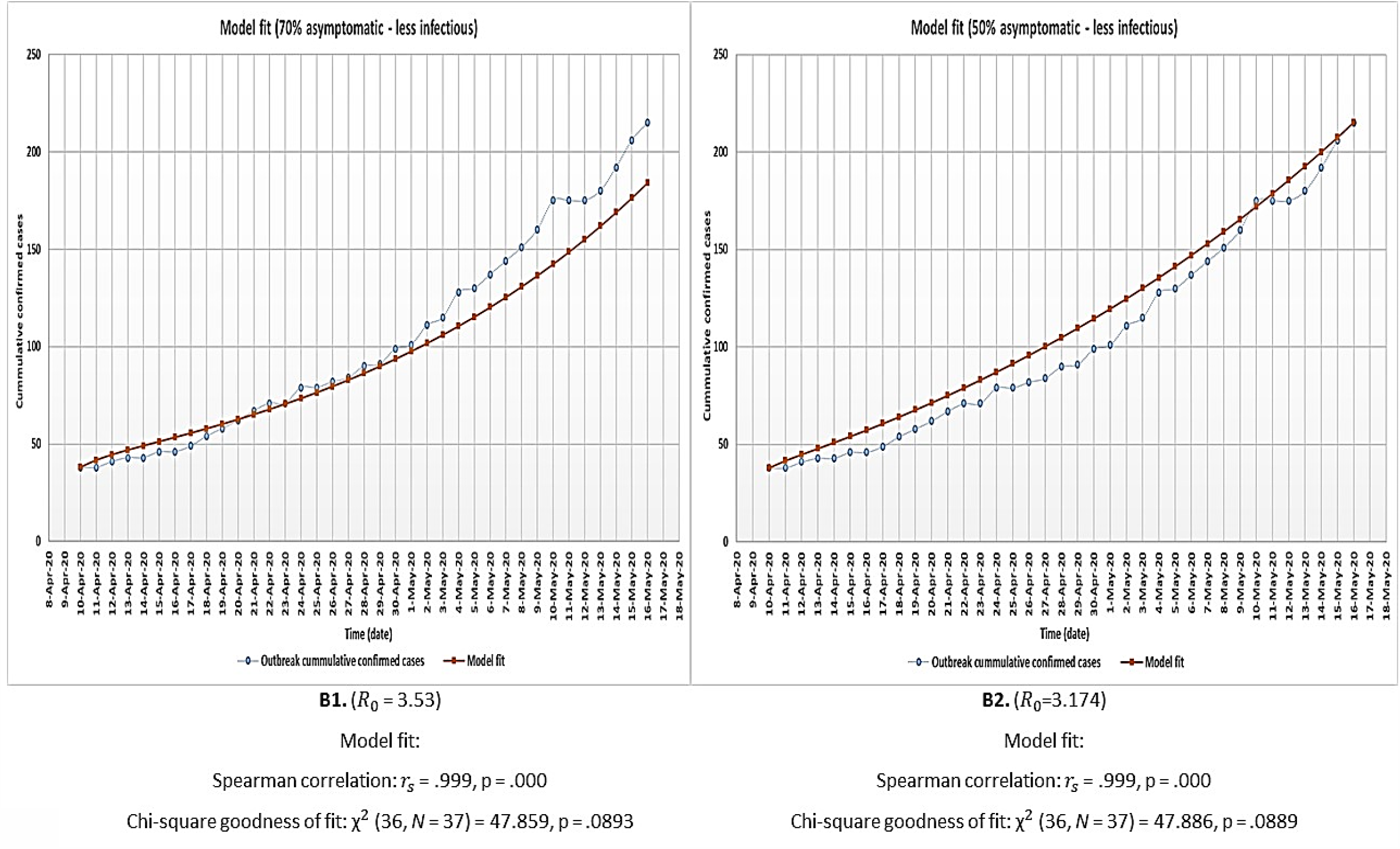
Model optimisation and fit to outbreak data for both the 70% (B1) and 50% (B2) proportion asymptomatic individual’s scenarios under the assumption that asymptomatic individuals are only half as infectious as symptomatic individuals

At both assessed levels of infectiousness for asymptomatic individuals, Table 2 shows that the model rightly predicts slightly lower expected peak values for the cumulative number of confirmed cases if there is more (70%) of asymptomatic individuals in the community who are poorly detected. However, this more substantial proportion of asymptomatic individuals (70%) is associated with higher expected peak values of both total active infectious individuals and total quarantined individuals. It is also observed that the outbreak is expected to peak much earlier for all variables in Table 2 for the 70% asymptomatic individual’s scenario compared to only 50% asymptomatic. Note that the model predictions for the Lusaka COVID-19 outbreak given in Table 2 are however subject to the effectiveness of the containment policies in Lusaka province over time. Figures 4 – 7 and Table 3 give the results of the effect of increasing blanket testing rates (θ) on control of the outbreak (flattening the curve for the expected cumulative number of confirmed cases and expected total number of active infectious individuals over time) for the assessed model scenarios. If asymptomatic individuals make up 70% of all COVID-19 cases and we assume equal infectiousness for asymptomatic and symptomatic individuals, then a minimum blanket testing rate of about ≥10028/100000 would be sufficient to flatten the curve for both the expected cumulative number of confirmed cases (Fig4 A1.1) and the total number of active infectious individuals (Fig4 A1.2) by more than 90% (Table 3). However, if the asymptomatic individuals only make up 50% of all cases and are also as infectious as symptomatic individuals, then a minimum blanket testing rate of ≥4540/100000 would be sufficient to flatten the curve for both the expected cumulative number of confirmed cases (Fig5 A2.1) and the total number of active infectious individuals (Fig5 A2.2) by more than 90% (Table 3). Alternatively, if asymptomatic individuals make up 70% of all COVID-19 cases but are only half as infectious as symptomatic individuals, then a minimum blanket testing rate of about ≥ 7911/100000 would be sufficient to flatten the curve for both the expected cumulative number of confirmed cases (Fig6 B1.1) and the total number of active infectious individuals (Fig6 B1.2) by more than 90% (Table 3). However, if the asymptomatic individuals only make up 50% of all cases and are only half as infectious relative to the symptomatic individuals, then a minimum blanket testing rate of ≥ 4383/100000 would be sufficient to flatten the curve for both the expected cumulative number of confirmed cases (Fig7 B2.1) and the total number of active infectious individuals (Fig7 B2.2) by more than 90% (Table 3) which is comparable to the result for A2.1 and A2.2.

**Table 2.**
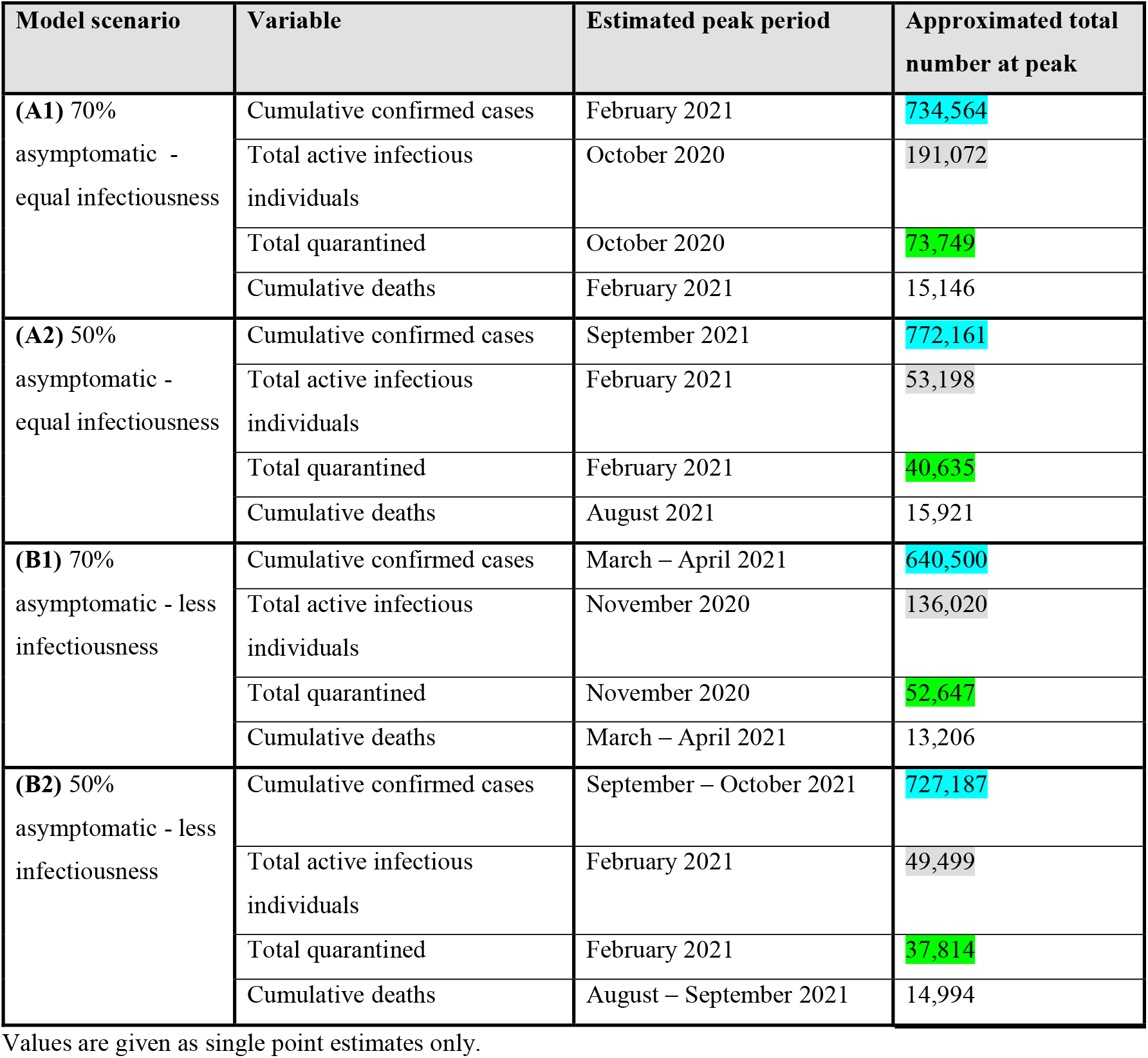
The model predicted key outbreak variables for the Lusaka COVID-19 outbreak for period 10^th^ April 2020 – 31^st^ December 2021 under the different model assumptions.

| Model scenario | Variable | Estimated peak period | Approximated total number at peak |
| --- | --- | --- | --- |
| (A1) 70% asymptomatic - equal infectiousness | Cumulative confirmed cases | February 2021 | 734,564 |
|  | Total active infectious individuals | October 2020 | 191,072 |
|  | Total quarantined | October 2020 | 73,749 |
|  | Cumulative deaths | February 2021 | 15,146 |
| (A2) 50% asymptomatic - equal infectiousness | Cumulative confirmed cases | September 2021 | 772,161 |
|  | Total active infectious individuals | February 2021 | 53,198 |
|  | Total quarantined | February 2021 | 40,635 |
|  | Cumulative deaths | August 2021 | 15,921 |
| (B1) 70% asymptomatic - less infectiousness | Cumulative confirmed cases | March – April 2021 | 640,500 |
|  | Total active infectious individuals | November 2020 | 136,020 |
|  | Total quarantined | November 2020 | 52,647 |
|  | Cumulative deaths | March – April 2021 | 13,206 |
| (B2) 50% asymptomatic - less infectiousness | Cumulative confirmed cases | September – October 2021 | 727,187 |
|  | Total active infectious individuals | February 2021 | 49,499 |
|  | Total quarantined | February 2021 | 37,814 |
|  | Cumulative deaths | August – September 2021 | 14,994 |

Values are given as single point estimates only.

**Table 3.** Approximated percentage reduction in the predicted cumulative number of confirmed cases and total number of active infectious individuals for higher values of θ (blanket testing rate).

| Model scenario | Variable | $\theta$ (tests/100000 populations) | percent reduction |
| --- | --- | --- | --- |
| **(A1)** 70% asymptomatic - equal infectiousness ($R_0 = 2.23$) | Cumulative confirmed cases (A1.1) | 9322/100000 | $\approx 77.27\%$ |
| | | 10028/100000 | $\approx 92.98\%$ |
| | | 10733/100000 | $\approx 98.12\%$ |
| | Total active infectious individuals (A1.2) | 9322/100000 | $\approx 96.92\%$ |
| | | 10028/100000 | $\approx 98.99\%$ |
| | | 10733/100000 | $\approx 99.81\%$ |
| **(A2)** 50% asymptomatic - equal infectiousness ($R_0 = 2.03$) | Cumulative confirmed cases (A2.1) | 3991/100000 | $\approx 88.26\%$ |
| | | 4540/100000 | $\approx 96.175\%$ |
| | | 5089/100000 | $\approx 98.74\%$ |
| | Total active infectious individuals (A2.2) | 3991/100000 | $\approx 92.76\%$ |
| | | 4540/100000 | $\approx 98.04\%$ |
| | | 5089/100000 | $\approx 99.58\%$ |
| **(B1)** 70% asymptomatic - less infectiousness ($R_0 = 3.53$) | Cumulative confirmed cases (B1.1) | 7206/100000 | $\approx 75.42\%$ |
| | | 7911/100000 | $\approx 92.44\%$ |
| | | 8617/100000 | $\approx 97.998\%$ |
| | Total active infectious individuals (B1.1) | 7206/100000 | $\approx 95.18\%$ |
| | | 7911/100000 | $\approx 98.44\%$ |
| | | 8617/100000 | $\approx 99.72\%$ |
| **(B2)** 50% asymptomatic - less infectiousness ($R_0 = 3.174$) | Cumulative confirmed cases (B2.1) | 3678/100000 | $\approx 72.86\%$ |
| | | 4383/100000 | $\approx 91.55\%$ |
| | | 5089/100000 | $\approx 97.82\%$ |
| | Total active infectious individuals (B2.2) | 3678/100000 | $\approx 85.66\%$ |
| | | 4383/100000 | $\approx 95.03\%$ |
| | | 5089/100000 | $\approx 99.101\%$ |

**Figure 4.**
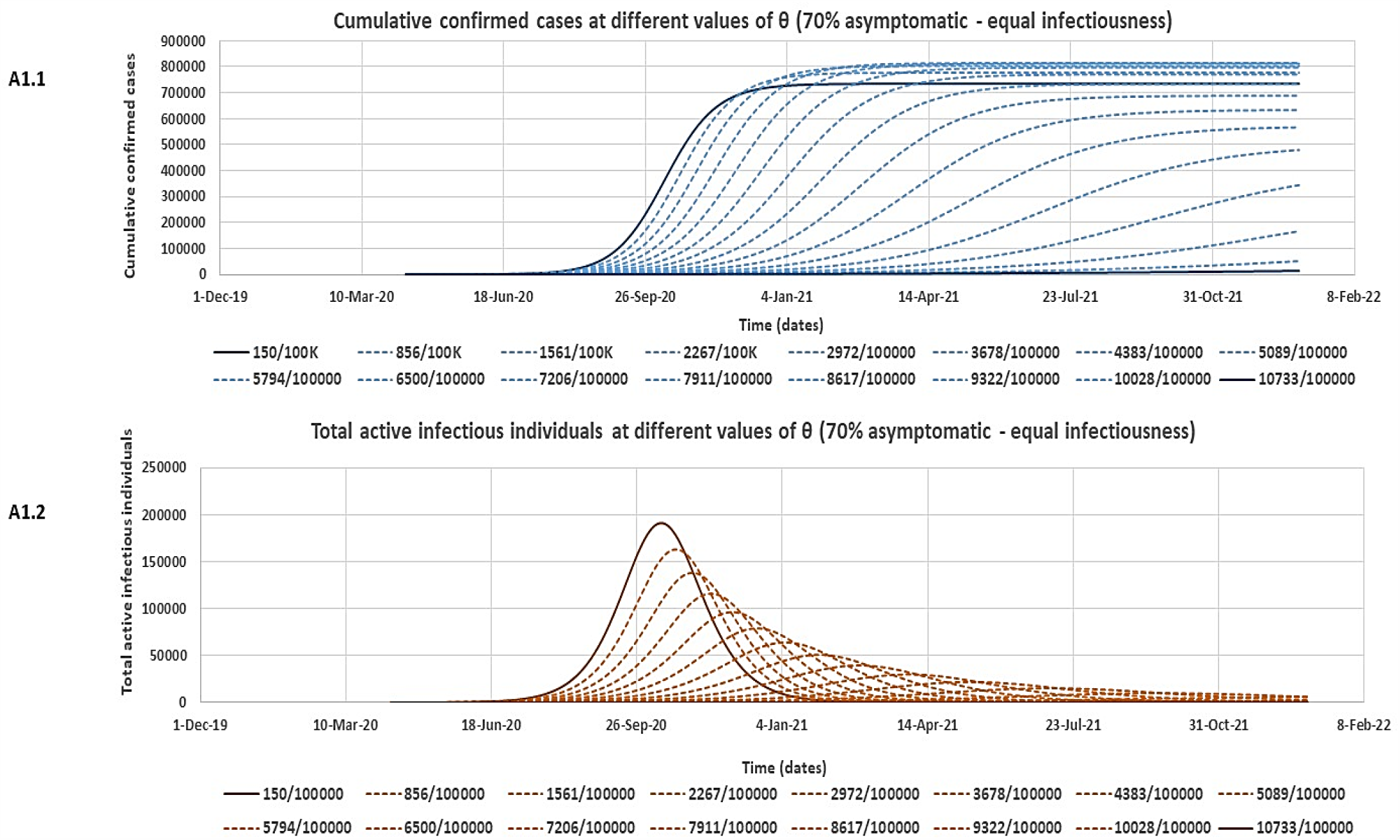
The estimated reduction in the cumulative number of confirmed cases (A1.1) and the total number of active infectious individuals (A1.2) at different values of θ (blanket testing rate) for the 70% asymptomatic scenario assuming equal infectiousness for asymptomatic and symptomatic individuals.

**Figure 5.**
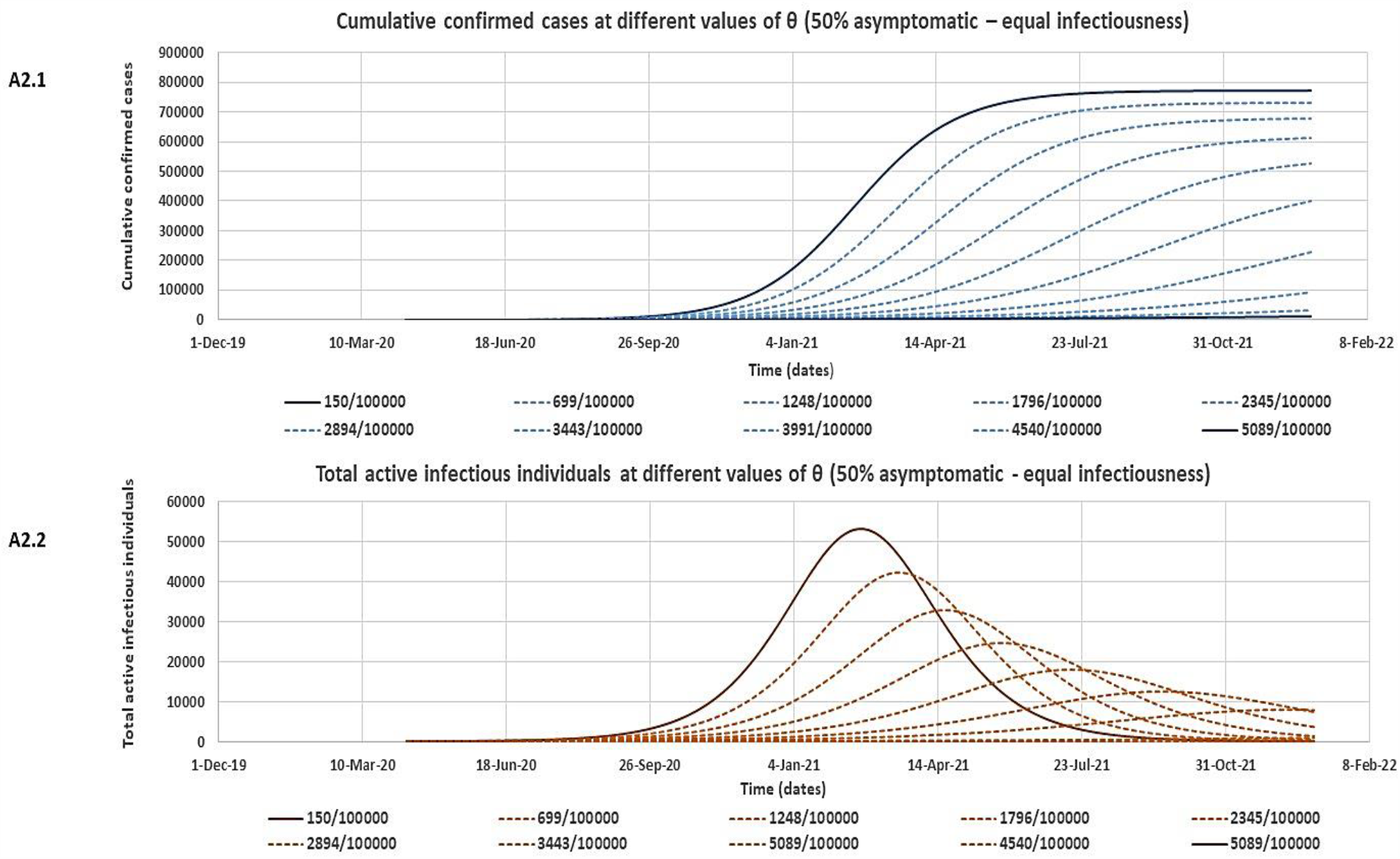
The estimated reduction in the cumulative number of confirmed cases (A2.1) and the total number of active infectious individuals (A2.2) at different values of θ (blanket testing rate) for the 50% asymptomatic scenario assuming equal infectiousness for asymptomatic and symptomatic individuals.

**Figure 6.**
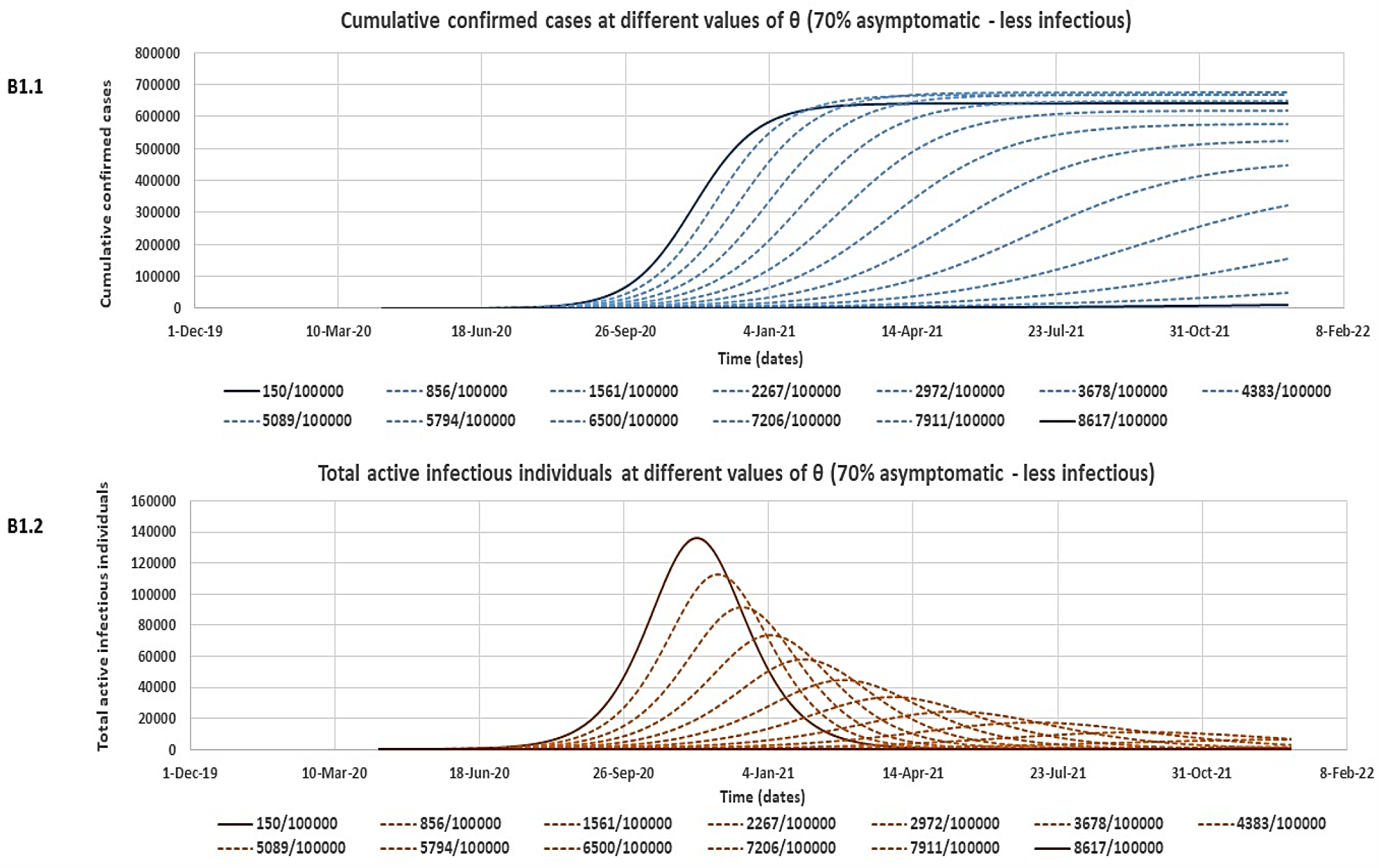
The estimated reduction in the cumulative number of confirmed cases (B1.1) and the total number of active infectious individuals (B1.2) at different values of θ (blanket testing rate) for the 70% asymptomatic scenario assuming asymptomatic individuals are only half as infectious as symptomatic individuals.

**Figure 7.**
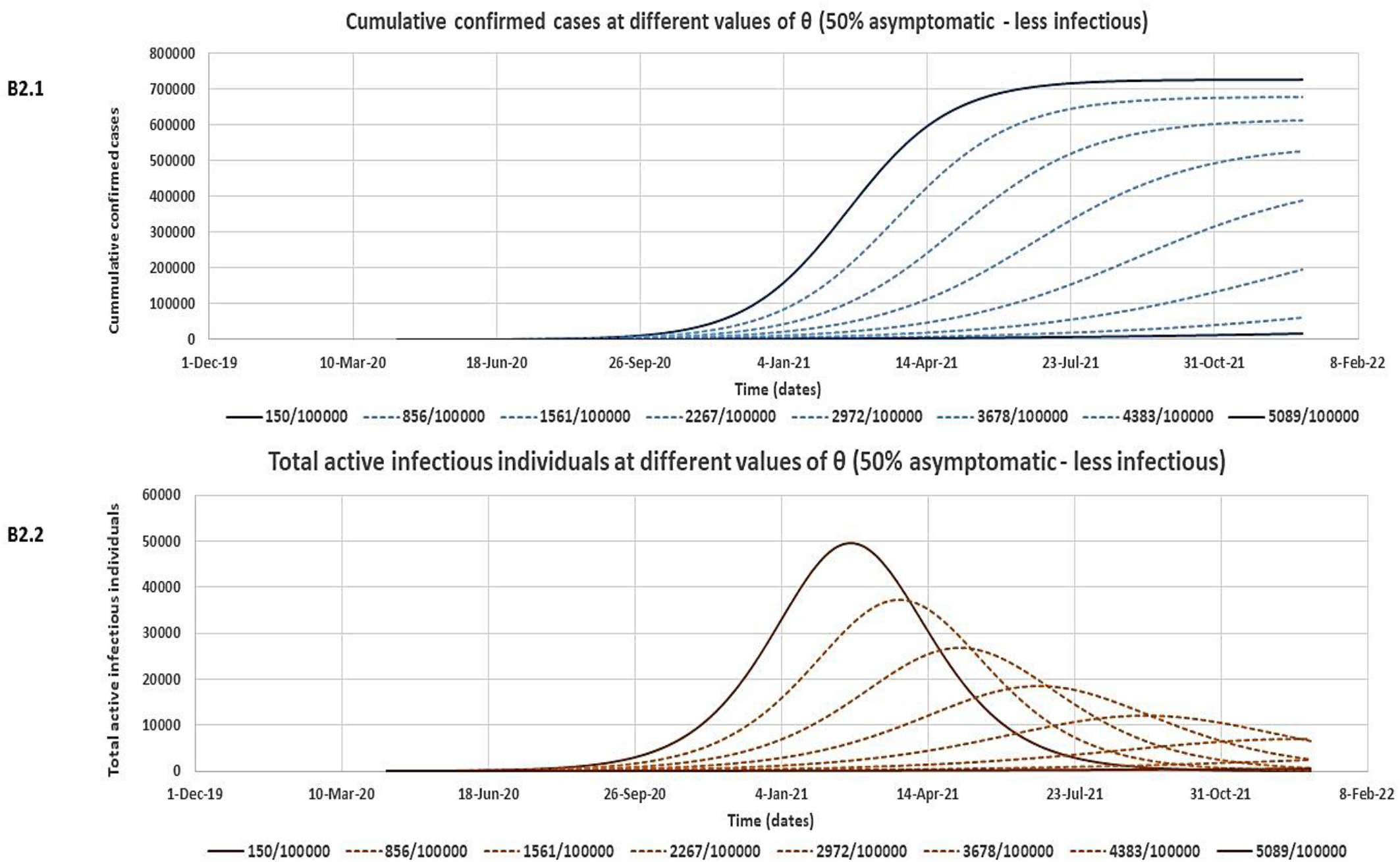
The estimated reduction in the cumulative number of confirmed cases (B2.1) and the total number of active infectious individuals (B2.2) at different values of θ (blanket testing rate) for the 50% asymptomatic scenario assuming asymptomatic individuals are only half as infectious as symptomatic individuals

## Discussion

In this study, we developed a simple deterministic model to forecast the spread of infection and assess required blanket testing rates for the control of the novel SARS-Cov-2 outbreak in Lusaka province, Zambia, with specific consideration for asymptomatic infectious individuals. In the early stages of novel infectious diseases, stochastic models are used due to uncertainty in parameter values. However, deterministic models which require average values can be used after the pandemic has progressed as is the case in this study and other studies that have used deterministic models to study COVID-19 [17,64]. In the context of Africa, earlier modelling studies have given gloomy predictions of the pandemic for the continent in case of failed early containment [42,43,50,51,53] with Zambia projected to have more than 2.8 million total infections [53] or between 4.8 million and 5.7 million total symptomatic cases [43] in the first year with estimated outbreak peaks as early as July 2020 [42,43, 53]. Further, the model in [43] also predicted that Zambia would record between 2.5 million – 4.4 million total number of symptomatic cases over a 12 months even under various levels of physical distancing and shielding interventions applied at country level. However, early implementation of the outbreak containment measures in Zambia, and most African countries, appears to be substantially averting the given predictions [53,44]. This may be because Africa was on high alert and prepared for the virus given the weaker health systems on the continent hence early containment of the disease significantly slowed the pandemic. Further, other studies have pointed to favourable bio-socioecological factors on the African continent leading to the lower rate of transmission in the region with fewer expected cases and deaths compared to other countries such as USA and Italy [53,64,65]. With limited incorporation of current containment measures implemented in Zambia, the highest number of cumulative confirmed cases predicted for Lusaka province in our model was 772,161 cases by September 2021 (Table 2). This lower projection is plausible given that it is an estimate for Lusaka province alone and it conforms to the relatively slow progression of the outbreak in Zambia compared to earlier predictions [44,53,64,65]. Further, our model predicts that the outbreak in Lusaka would spread much faster and peak earlier if asymptomatic individuals make up 70% of all COVID-19 cases (earliest peaks October - November 2020 for total active infectious individuals). This is because a higher proportion of poorly detected asymptomatic individuals inflated the population of active infectious individuals in the population (*I*_*S*_ + *I*_*A*_ = *I*_*T*_) thereby increasing the infection rates in the community; 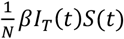 (ODEs in set-1) or 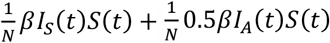 (ODEs in set-2), resulting in a faster progression of the outbreak. The higher proportion of asymptomatic individuals was, however, associated with less overall cumulative number of confirmed cases since asymptomatic individuals are generally less diagnosed under targeted testing approaches such as those employed in Lusaka province [53].

In our model, we refer as asymptomatic all “silent spreaders” of COVID-19 which includes all those that have become infectious but never develop symptoms (asymptomatic or mildly symptomatic) and those that have become infectious but only develop symptoms much later in their infection (pre-symptomatic spreaders) [53,55-59]. Studies however agree that collectively these asymptomatic individuals (or silent spreaders) are less infectious compared to symptomatic individuals [57,61,62]. This is most likely because their viral shedding rate (how much virus an infected person releases) may be limited without the symptoms of coughing and sneezing which produces infectious respiratory droplets even if these asymptomatic individuals may have similar viral load, duration of viral shedding, and contact rates in the community as symptomatic individuals [2,60]. Therefore, we suspect that transmission of the infection from asymptomatic individuals may mostly rely on the other possible routes of transmission for COVID-19 including contact with fomites, unwitting close intimate facial contact, and possibly even fecal-oral route [53,62,63]. Even with a lowered ability to transmit the infection, lack of detection and awareness of the carrier state may cause the asymptomatic individuals to be playing an important role in the spread of COVID-19 and sustaining the outbreak [2,3,4,24,25,33]. Active isolation of all infected individuals (symptomatic and asymptomatic) through blanket testing therefore offers a possible solution to this challenge [3,4,53]. In our study, we assessed the effect of various levels of blanket testing for control of the COVID-19 outbreak in Lusaka province under different assumed scenarios of the proportion and infectiousness of asymptomatic individuals with results tabulated in Figures 4 – 7 and Table 3. Out of all the assessed scenarios, we nominate scenario B1 to represent the most likely epidemiological dynamics for COVID-19 in the population where asymptomatic individuals make up 70% of cases but are only half as infectious as symptomatic individuals. Therefore, for this scenario, a blanket testing rate of about ≥ 7911 tests /100000 (≈ 80/1000) would be sufficient to control the outbreak in Lusaka. To compare with other possible outbreak control strategies, note that the model fitted value of R_0_ for the selected scenario B1 was 3.53 (Figure 3 B1). Therefore, in case of available vaccine for COVID-19, a minimum protective vaccination rate of about 1 − 1/*R*_0_ (≈70%) would be required to control the outbreak in the population; assuming vaccination provides full protection [66-69]. This proportionately translates into roughly 70000 vaccinations /100000 populations which is much higher than the estimated required blanket testing intervention. The effect of blanket testing is that it acts to directly reduce the size of the total number of active infectious individuals which leads to a disproportionately higher reduction in the infection rate in the population. This is achieved through active isolation of even the otherwise largely un-detected asymptomatic individuals in the population who possibly participate in the transmission of the infection [2,3,4,24]. In the case of Africa, control of asymptomatic infections may be even more pertinent than other regions of the world. This is because Africa has been found to have a younger population and with lower personal vulnerabilities to severe infection compared to Europe, USA, and other regions leading to most of the infections in Africa being asymptomatic [53,64,65]. Therefore, more outbreak control strategies with specific consideration of the role of asymptomatic individuals will be required in Africa.

## Conclusion

Active isolation of SARS-Cov-2 asymptomatic infectious individuals through blanket testing has been shown to have indeed the potential to control the outbreak in Lusaka province of Zambia. However, this can only be achieved at a rate of approximately ≥ 7911 tests /100000 (≈ 80/1000) following the most likely epidemiological characteristics of the COVID-19 outbreak in Lusaka.

## Limitations

As is the case in creating infectious disease models, appropriateness of the model design to incorporate containment measures implemented in Lusaka and quality of data used were some of the challenges faced in this study [70,71]. Also, limited outbreak data was used due to significant irregularities in the reported outbreak data. Cases were not reported in real-time and there is a possibility of under-reporting due to low testing capacities in the earlier stages of the outbreak in Zambia [44,54]. Additionally, the model was fitted to cumulative confirmed cases which could still introduce uncertainties [54]. Further modelling studies using more refined models and more outbreak data should, therefore, be conducted to study the COVID-19 outbreak in Lusaka.

## Data Availability

The data underlying the results presented in the study are available from the Zambia MOH/ZNPH/WHO, | New Coronavirus Disease of 2019 (COVID-19) Situation Reports [1-64]. March-May 2020. Accessed from http://znphi.co.zm/news/situation-reports-new-coronavirus-COVID-19-sitreps/.

## Abbreviations

COVID-19: Novel Coronavirus disease
SARS-CoV-2: Severe acute respiratory syndrome Coronavirus 2
CFR: Case fatality rate
SEIR: susceptible-exposed-infectious-removed compartmental mathematical model
ODEs: Ordinary differential equations

